# Diagnosis of celiac disease on a gluten-free diet: a multicenter prospective quasi-experimental clinical study

**DOI:** 10.1101/2024.08.05.24311406

**Authors:** Sara Gómez-Aguililla, Sergio Farrais, Natalia López-Palacios, Beatriz Arau, Carla Senosiain, María Corzo, Nora Fernandez-Jimenez, Ángela Ruiz-Carnicer, Fernando Fernández-Bañares, Bárbara P. González-García, Eva Tristán, Ana Montero-Calle, María Garranzo-Asensio, Isabel Casado, Mar Pujals, Juana María Hernández, Jorge Infante-Menéndez, Garbiñe Roy, Carolina Sousa, Concepción Núñez

## Abstract

**Background:** Diagnosing celiac disease (CD) in individuals adhering to a gluten-free diet (GFD) presents significant challenges. Current guidelines recommend a gluten challenge (GC) lasting at least 6-8 weeks, which has several limitations.

**Objectives:** This study compares four approaches previously proposed for diagnosing CD on a GFD: IL-2 serum levels, gut-homing CD8^+^ T cells, %TCRγδ^+^ intraepithelial lymphocytes (IELs), and *UBE2L3* gene expression. Additionally, we evaluated the CD8^+^ T-cell based method with a 3-day GC against the standard GC protocol.

**Methods:** We conducted a multicenter prospective quasi-experimental clinical study. Two subsets of individuals were considered: 1) 20 patients with CD and 15 non-CD controls previously diagnosed, to evaluate the first aim; 2) 45 individuals with uncertain diagnosis who were on a GFD and required GC following current clinical guidelines, to assess the second aim. All participants underwent a 3-day GC (10 g gluten/day).

**Results:** Among CD patients and non-CD controls, the sensitivity and specificity of IL-2, gut-homing CD8^+^ T cells, and *UBE2L3* were 82.4% and 83.3%, 88.2% and 100%, and 52.9% and 100%, respectively. The percentage of TCRγδ^+^ IELs showed 88.2% sensitivity. In the uncertain diagnosis group, a CD8^+^ T-cell positive response was observed in 8 of the 45 subjects.

**Conclusion:** The percentage of TCRγδ^+^ IELs and the gut-homing CD8^+^ T-cell assay are promising diagnostic methods for CD on a GFD. Notably, the CD8^+^ T-cell assay provides a consistent and reliable alternative to the extended GC, eliminating the need for the invasive procedures to obtain duodenal samples and the prolonged gluten ingestion.

## INTRODUCTION

Celiac disease (CD) is an immune-mediated enteropathy triggered by gluten ingestion in genetically predisposed individuals [1]. The diagnosis relies on the clinical suspicion driven by symptoms or risk factors, which initiates a diagnostic work-up including detection of specific serologic markers and histological alterations. In certain cases, HLA testing can be useful [2]. This established diagnostic strategy requires the tested individual to maintain a gluten-containing diet.

Diagnosing CD becomes challenging when patients have already initiated a gluten-free diet (GFD) without prior testing [3, 4] or in cases of discrepant serological and histological results. The gluten challenge (GC) is not routinely included in standard algorithms for CD diagnosis; nevertheless, it plays a crucial role in these cases of uncertain diagnosis. However, this is far from an optimal solution, because of two critical considerations. First, a considerable number of patients refuse this procedure owing to concerns about clinical relapse. Second, the absence of a standardized GC protocol introduces uncertainties regarding the appropriate amount of gluten to be ingested and the optimal duration of exposure. Current guidelines suggest different strategies for conducting GC, such as a regimen of at least 3 g/day for 2–8 weeks [3] or a higher intake of 10 g/day for 6–8 weeks [5].

In the past two decades, increasing knowledge of the pathogenesis of CD has opened new prospects for diagnosing CD while on a GFD. Methods based on the immunological response to a short GC have emerged as highly promising approaches [6, 7, 8, 9, 10]. In 2021, Leonard et al. conducted a preliminary study by comparing the changes observed in various previously described markers after a short GC using two gluten doses (3 g and 10 g gluten/day) [11]. Specifically, they evaluated the change in the ratio of villous height to crypt depth following a 14-day GC; HLA-DQ2-gliadin tetramer, IFN-γ ELISpot and gut-homing CD8^+^ T-cells at baseline and at day 6 after GC; and plasma IL-2 levels at baseline and 4 hours after a single gluten exposure. Preferential responses were observed in patients receiving 10 g of gluten, and IL-2 was described as the earliest and most sensitive marker. The short GC was well tolerated with minimal variation in clinical symptoms among individuals in both groups. The CD8^+^ T-cell assay stood out as a noteworthy approach in the comparison baseline-day 6 because it required the lowest volume of blood and did not necessitate *in vitro* culture or enrichment, making it a feasible technique in both research and clinical settings. A previous study by our group demonstrated that gut-homing CD8^+^ T cells provide accurate CD diagnosis with 95% specificity and 97% sensitivity [12].

Furthermore, other methods with no challenge requirements have been proposed for diagnosing CD on a GFD, including the increase in TCRγδ^+^ intraepithelial lymphocytes (IELs) determined by flow cytometry [13, 14, 15] or the relative expression of *UBE2L3* isoforms, an ubiquitin ligase located in a CD-associated region [16]. A comprehensive comparison of these methods with the previously described techniques is still warranted.

We aimed to evaluate the most appropriate method for diagnosing CD in patients on a GFD based on diagnostic accuracy and feasibility of potential translation into daily clinical practice. We considered four approaches previously proposed in the literature (serum IL-2 levels, gut-homing CD8^+^ T cells, percentage of TCRγδ^+^ IELs, and *UBE2L3* gene expression. We tested them in the same group of patients with a previously confirmed diagnosis. Additionally, we evaluated the CD8^+^ T-cell assay in a real-world scenario with a group of patients who had an uncertain diagnosis of CD and who required a gluten-containing diet for ≥ 6-8 weeks, in accordance with clinical guidelines. We then compared these results with those obtained after a 3-day GC.

## MATERIAL AND METHODS

### Study design

A multicenter prospective, quasi-experimental clinical study was conducted at four tertiary centers in Spain: Hospital Universitari Mutua Terrassa (Barcelona), Hospital Clínico San Carlos, Hospital Universitario Fundación Jiménez Díaz, and Hospital Universitario Ramón y Cajal (Madrid). The study protocol was approved by the ethical committees of the four participating hospitals: CEIm Hospital Clínico San Carlos, Madrid, Spain (December 22, 2020); Fundació Assistencial Mútua Terrassa, Barcelona, Spain (January 27, 2021); Fundación Jiménez Díaz, Madrid, Spain (March 9, 2021); Hospital Universitario Ramón y Cajal, Madrid, Spain (August 17, 2021). Informed consent was obtained from all the subjects.

### Participants

All patients were consecutively recruited during clinic visits at the participating hospitals. Only adult patients (≥ 18 years old) on a GFD ≥ 1 month were enrolled. The exclusion criteria included severe reactions to unintentional previous gluten transgressions in the past, recent positive anti-transglutaminase type 2 antibodies (ATG2), and ongoing immunosuppressive treatment.

Two distinct subsets were considered in the present study (Figure 1). The first group consisted of 35 subjects recruited from July 2021 to July 2023 to establish the diagnostic value of different tests: 1) 20 patients previously diagnosed with serology and biopsy consistent with CD, and 2) 15 non-CD controls. This included 9 healthy controls recruited from personal staff, and their relatives, or friends, without known medical conditions or symptoms suggestive of CD, and 6 patients with suspected gluten-related functional bowel disease symptoms (SGRS) and normal duodenal mucosa at diagnosis. All controls were negative for ATG2 while maintaining a gluten-containing diet. Healthy controls adhered to a strict GFD the month prior to the study, with dietary compliance monitored by gluten immunogenic peptides (GIP). The second subset of participants included 45 individuals with an uncertain diagnosis due to a self-prescribed GFD, incomplete testing, or equivocal serological/histological findings, who required GC according to current clinical guidelines. Demographic and clinical data were collected from all participants (Table 1).

**Figure 1.**
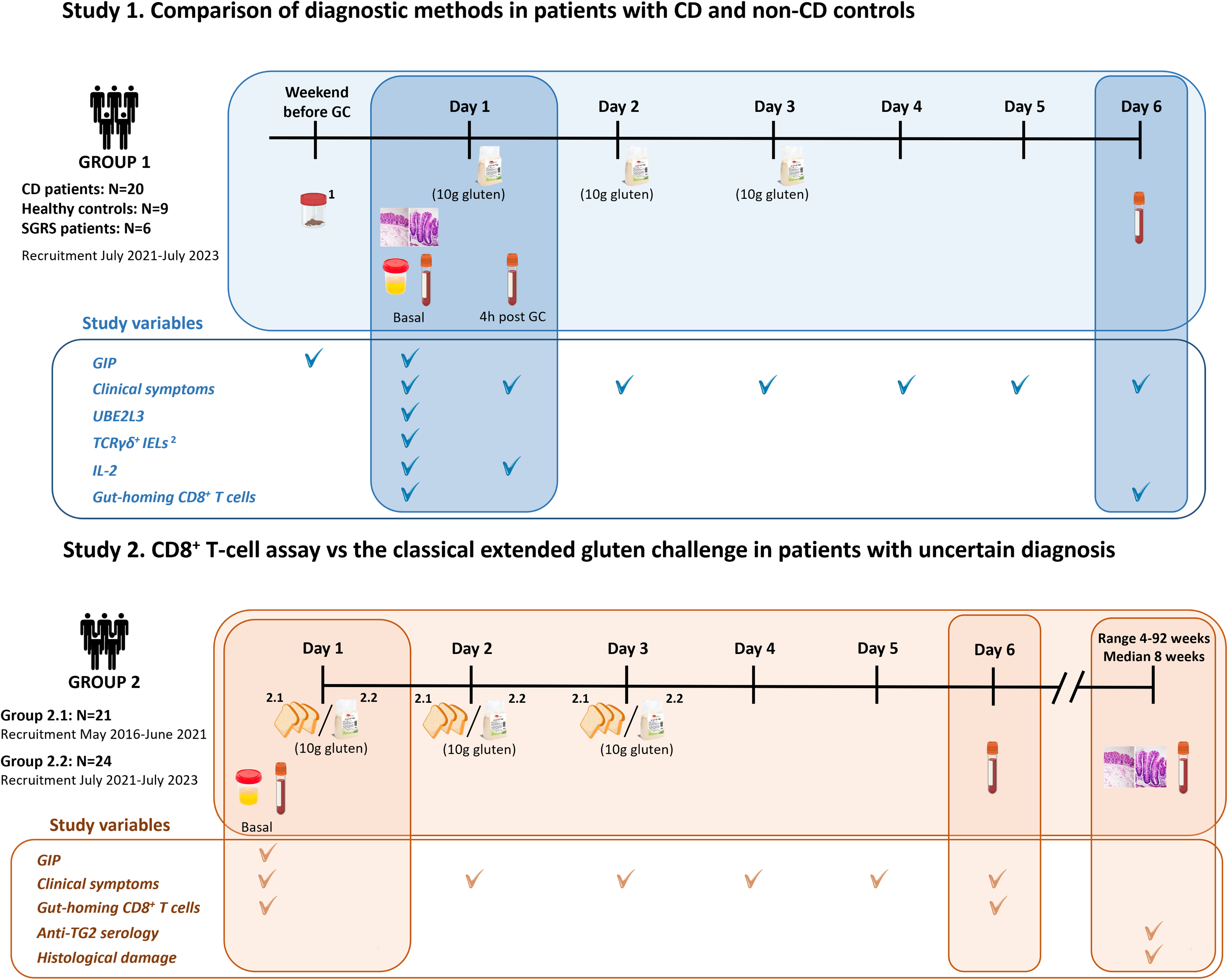
Design of the two studies presented. Number and type of participants, along with the timing of sample collection and the type of analysis performed are included. ^1^In patients with CD and SGRS. ^2^In patients with CD. 4h: 4 hours; anti-TG2: anti-transglutaminase type 2; CD: celiac disease; GC: gluten challenge; GIP: gluten immunogenic peptides; IELs: intraepithelial lymphocytes; SGRS: suspected gluten-related functional bowel disease symptoms.

**Table 1.**
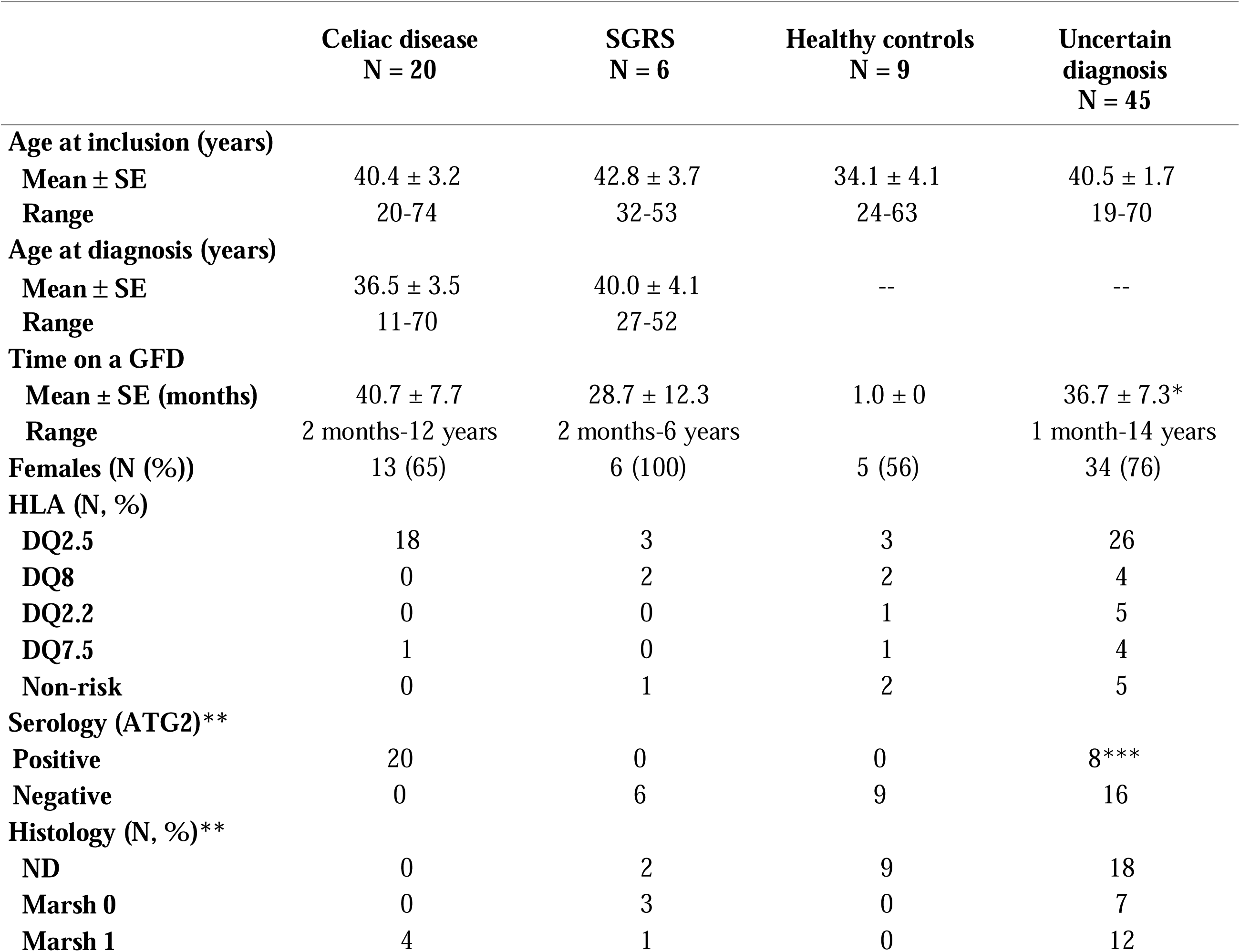
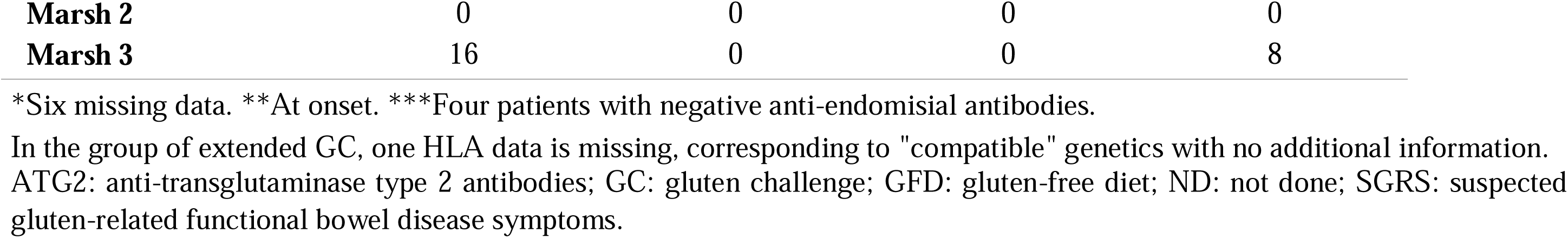
Characteristics of the participants in the study.

|  | **Celiac disease**<br>**N = 20** | **SGRS**<br>**N = 6** | **Healthy controls**<br>**N = 9** | **Uncertain**<br>**diagnosis**<br>**N = 45** |
| --- | --- | --- | --- | --- |
| **Age at inclusion (years)** |  |  |  |  |
| Mean ± SE | 40.4 ± 3.2 | 42.8 ± 3.7 | 34.1 ± 4.1 | 40.5 ± 1.7 |
| Range | 20-74 | 32-53 | 24-63 | 19-70 |
| **Age at diagnosis (years)** |  |  |  |  |
| Mean ± SE | 36.5 ± 3.5 | 40.0 ± 4.1 | -- | -- |
| Range | 11-70 | 27-52 |  |  |
| **Time on a GFD** |  |  |  |  |
| Mean ± SE (months) | 40.7 ± 7.7 | 28.7 ± 12.3 | 1.0 ± 0 | 36.7 ± 7.3* |
| Range | 2 months-12 years | 2 months-6 years |  | 1 month-14 years |
| **Females (N (%))** | 13 (65) | 6 (100) | 5 (56) | 34 (76) |
| **HLA (N, %)** |  |  |  |  |
| DQ2.5 | 18 | 3 | 3 | 26 |
| DQ8 | 0 | 2 | 2 | 4 |
| DQ2.2 | 0 | 0 | 1 | 5 |
| DQ7.5 | 1 | 0 | 1 | 4 |
| Non-risk | 0 | 1 | 2 | 5 |
| **Serology (ATG2)\*\*** |  |  |  |  |
| Positive | 20 | 0 | 0 | 8*** |
| Negative | 0 | 6 | 9 | 16 |
| **Histology (N, %)\*\*** |  |  |  |  |
| ND | 0 | 2 | 9 | 18 |
| Marsh 0 | 0 | 3 | 0 | 7 |
| Marsh 1 | 4 | 1 | 0 | 12 |
| **Marsh 2** | 0 | 0 | 0 | 0 |
| **Marsh 3** | 16 | 0 | 0 | 8 |

\*Six missing data. \*\*At onset. \*\*\*Four patients with negative anti-endomisial antibodies.
In the group of extended GC, one HLA data is missing, corresponding to "compatible" genetics with no additional information. ATG2: anti-transglutaminase type 2 antibodies; GC: gluten challenge; GFD: gluten-free diet; ND: not done; SGRS: suspected gluten-related functional bowel disease symptoms.

### Intervention and sample collection

All participants underwent a 3-day GC consisting of 1) 160 g of gluten-containing sliced white bread (approximately 10 g of gluten) administered daily in the morning from day 1 to day 3 or 2) 10 g of low FODMAP powdered gluten (El Granero Integral™; Biogran S.L., Madrid, Spain) administered daily in a lactose-free liquid yogurt between 8 and 11 a.m. from day 1 to day 3. This second schedule was the adopted for subjects recruited after July 2021 for standardization of GC for use in clinical settings. All participants completed the 3-day GC except for one patient with CD who discontinued treatment on day 2 owing to acute brain fog. The first morning urine sample was collected from all patients on the day of the intervention to assess dietary compliance with the GIP [17]. Additionally, stool samples were collected from patients with CD and SGRS on weekend before gluten reintroduction for the same purpose.

In the first subset of patients, upper endoscopy with duodenal biopsy was performed at baseline (prior to GC) in all patients with CD but not in controls. In all cases, peripheral blood was drawn before gluten ingestion for baseline measurements. Blood samples were additionally collected at 4 hours and 6 days after the first dose of gluten. Sample processing and analysis were blinded to the final diagnosis.

In the second subset, all participants underwent a 3-day GC with blood extraction at baseline and on day 6 to study the activated gut-homing CD8^+^ T-cells. Subsequently, they were recommended to maintain a more standard GC for at least 6-8 weeks followed by a duodenal biopsy and ATG2 serology. In two patients the period was reduced to 4 weeks due to clinical discomfort. The median duration of GC was 8 weeks (Q1-Q3 quartiles: 7-15 weeks). At the end of the follow-up period, duodenal biopsy was performed in all except three patients with non-compatible HLA genetics, and ATG2 was unavailable in six patients.

### *UBE2L3* gene expression

RNA was extracted from peripheral blood mononuclear cells (PBMC), previously isolated by density-gradient centrifugation and preserved in Nucleoprotect at 4 °C, using the NucleoSpin RNA kit (Macherey-Nagel GmbH & Co. KG, Dueren, Germany). Exon 4 and exon 5 of *UBE2L3* were quantified using the TaqMan®-based one-step RT-qPCR protocol. Exon 4 is specific to the non-coding *UBE2L3* isoform 2, whereas exon 5 is common to all annotated *UBE2L3* isoforms. To design these expression assays, first, *UBE2L3* exon 4 and exon 5 sequences were downloaded from GeneBank (NR_028436 accession number). Subsequently, we designed primers and probes for the TaqMan® expression assays using the “Custom TaqMan® Assay Design Tool” option on the ThermoFisher Scientific website. After bioinformatics analysis aimed at finding the most suitable expression assays, a custom assay for *UBE2L3* exon 4 and a commercially available assay for *UBE2L3* exon 5 were selected.

All samples were analyzed in triplicate with each TaqMan® assay processed in separate reactions. Ct was examined using Bio-Rad CFX Maestro software, and the ratio of Ct exon 4 to Ct exon 5 was determined.

### TCR**γδ**^+^ intraepithelial lymphocytes by flow cytometry

A single biopsy from the second part of the duodenum obtained before the 3-day GC was processed to obtain single-cell suspensions from the epithelial layer, which were stained and analyzed by flow cytometry as previously described [18]. Sample processing started within the first 4 hours after sample collection. Since samples correspond to patients on a GFD, the isolated increase of TCRγδ^+^ IELs ≥ 14% was considered for diagnosis [14].

### IL-2 levels

IL-2 was quantified in serum samples collected at baseline and 4 hours after the first dose of gluten using the single molecule counting (SMC™) human IL-2 high sensitivity immunoassay kit (EMD Millipore Corporation, US), which showed a lower limit of quantification (LLOQ) of 0.09 pg/mL. According to the literature, IL-2 serum levels ≥ 0.5 pg/mL at 4 hours were considered as a positive result [19], provided that lower levels were observed at baseline (post-4 hour/baseline ratio ≥ 1.7).

### Activated gut-homing CD8^+^ T cells by flow cytometry

Basal and post-6 day blood samples were stained and analyzed using conventional flow cytometry as previously detailed [12, 20]. Data analysis was performed upon completion of patient recruitment using the Kaluza Analysis Software (Beckman Coulter, CA, USA) with Batch Processing. In most patients recruited from July 2021, the CD8^+^ T cell population was characterized by expression of the previously described markers CD8^+^ CD103^+^ β7^hi^ CD38^+^ along with the addition of CCR9^+^. Using sliced bread for GC, we have previously described accurate CD diagnosis based on a proportion of activated gut-homing CD8^+^ T cells to total CD8^+^ T cells at day 6 ≥ 0.01%, contingent upon the presence of a ratio day 6/day 0 ≥ 2 [12].

### Clinical response

The clinical response to the GC was uniformly evaluated in all subjects in the first subset and in 24 patients in the second subset (those recruited from July 2021). It was assessed using a 9-item visual analog scale (VAS) ranging from no symptoms to severe symptoms. Flatulence, abdominal distension, abdominal pain, diarrhea, nausea, vomiting, tiredness, irritability, and brain fog were considered. The patients were instructed to document their symptoms daily from the day before the study to day 6. On day 1, symptom severity was documented 4 hours after gluten ingestion. The total symptoms severity ranged from 9 (no symptoms) to 45 (severe in the 9 recorded items).

### GIP test

Quantification of GIP in the stool samples was performed by enzyme-linked immunosorbent assay (ELISA) (iVYLISA GIP Stool, Biomedal S.L., Seville, Spain) following the manufacturer’s instructions as previously described [21]. The quantification limit of GIP was 0.078 μg/g of stool. The samples were tested in duplicate on two different days. The absence of GIP in the urine samples was verified using a lateral flow immunochromatographic assay (iVYCHECK GIP Urine, Biomedal S.L., Seville, Spain) [22, 23]. The GIP are resistant to gastrointestinal digestion and account for immunogenic reactions in the T cells of patients with CD [21, 22, 24].

### Statistical analysis

The optimal cut-off points for diagnosing CD based on the percentage of gut-homing CD8^+^ T cells after a 3-day GC using powdered gluten and the *UBE2L3* Ct exon 4/Ct exon 5 ratio were determined through Receiver Operating Characteristic (ROC) curve analysis using SPSS version 15.0.

Differences in the *UBE2L3* Ct exon 4/Ct exon 5 ratio between the CD and non-CD groups were evaluated using Student’s t-test. Comparisons within each category of patients at different times (baseline vs. post-4 hour for IL-2, baseline vs. post-6 day for gut-homing CD8^+^ T cells, or baseline vs. days 1–6 for clinical symptoms) were performed using a one-sided Wilcoxon signed-rank test. The correlation between the different diagnostic markers (*UBE2L3* Ct exon 4/ Ct exon 5 ratio, percentage TCRγδ^+^ IELs, post-4 hour IL-2 levels, and percentage post-6 day gut-homing CD8^+^ T cells) was calculated using Spearman’s rank correlation after excluding participants who did not show an increase in IL-2 or gut-homing CD8^+^ T cells at 4 hours or at day 6, respectively.

Graphs were generated with R version 4.0.3 using the ggplot2 package and GraphPad Prism version 5.01.

Our primary endpoint was to compare the diagnostic accuracy of previously proposed methods for diagnosing CD in patients on a GFD. According to the literature, the four studied diagnostic approaches were observed in ≥ 90% of patients with CD and ≤ 10% of non-CD individuals. Considering these figures and accepting an alpha risk of 0.05, a statistical power of 80% was achieved with 9 subjects per group. Additionally, we established a secondary endpoint to assess the performance of activated gut-homing CD8^+^ T cells with short GC compared to a longer GC, as recommended in current clinical practice, to identify CD in patients on a GFD with an uncertain diagnosis.

## RESULTS

Eighty consecutive patients, 72.5% females, were included within the study period. Among them, 6 (3 CD and 3 SGRS patients) showed a positive GIP stool determination and were excluded from the calculation of diagnostic accuracy, but their results were examined to understand the importance of dietary compliance in the different diagnostic approaches.

### Comparison of diagnostic methods in patients with CD and non-CD controls

#### UBE2L3 gene expression

It was assessed in 19 patients with CD and 13 non-CD subjects. The *UBE2L3* Ct exon 4/Ct exon 5 ratio was significantly increased in the CD group compared to non-CD: 0.996 ± 0.009 vs. 0.960 ± 0.009, respectively (p=0.010). The calculated AUC was 0.799 (95% CI 0.638-0.960) and a value of 0.9944 was selected as the cut-off, yielding a sensitivity and specificity for CD diagnosis of 52.9% (9/17) and 100% (0/12), respectively (Figure 2). A negative diagnostic result for *UBE2L3* was observed in the two patients with CD evaluated for this marker who tested positive for fecal GIP.

**Figure 2.**
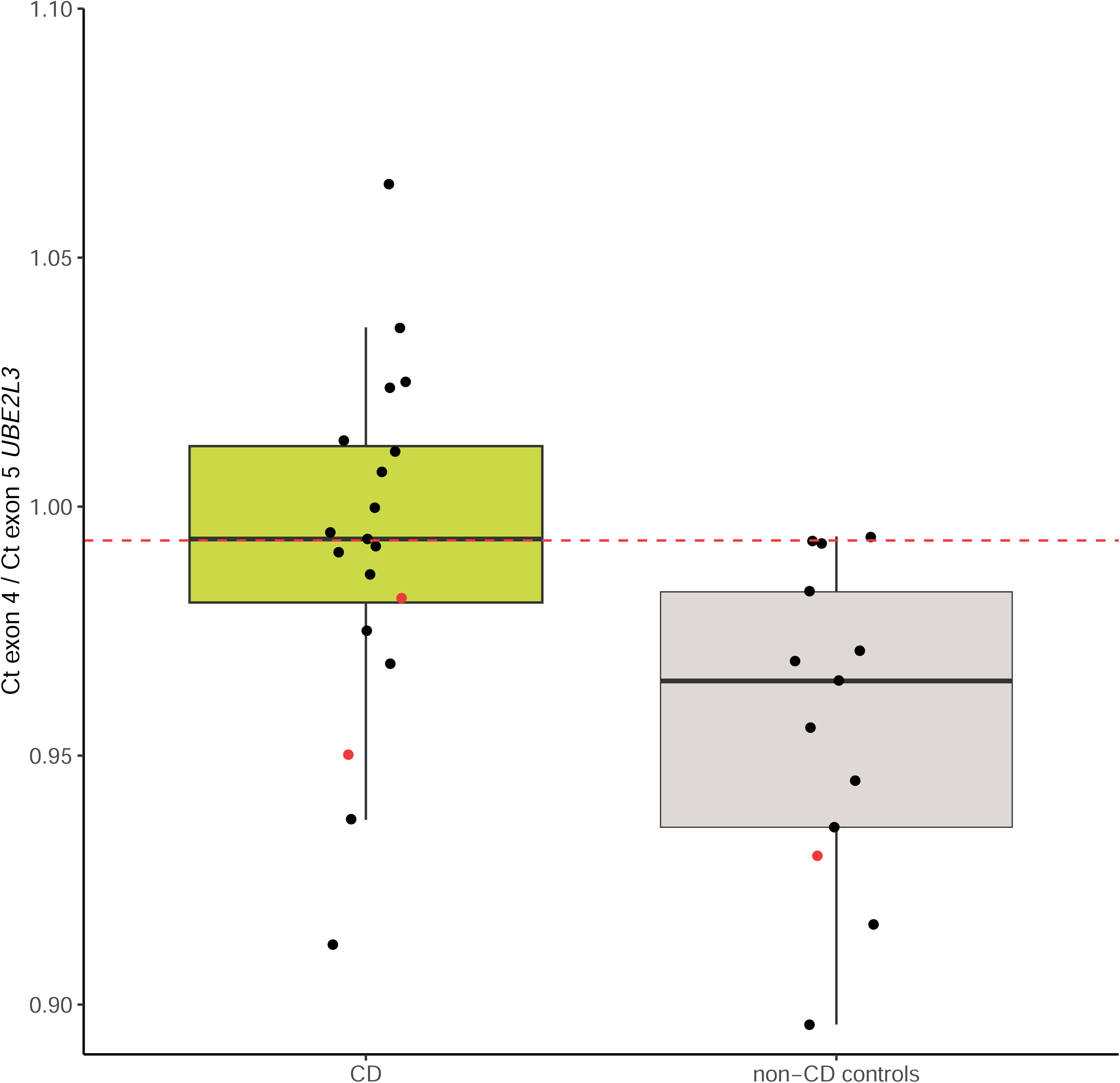
Baseline *UBE2L3* gene expression data. *UBE2L3* Ct exon 4 / Ct exon 5 ratio in subjects from each study group. The dotted red line indicates the cut-off for CD diagnosis. Red dots represent subjects with positive fecal GIP. Centerlines in the box represent the median; box limits indicate the 25th and 75th percentiles (IQR).

#### TCRγδ^+^ intraepithelial lymphocytes

This study included the 20 patients with CD. The isolated increase of TCRγδ^+^ IELs (≥ 14%) offered 88.2% (15/17) sensitivity (Figure 3). The three patients with CD with positive fecal GIP showed ≥ 14% TCRγδ^+^ IELs.

**Figure 3.**
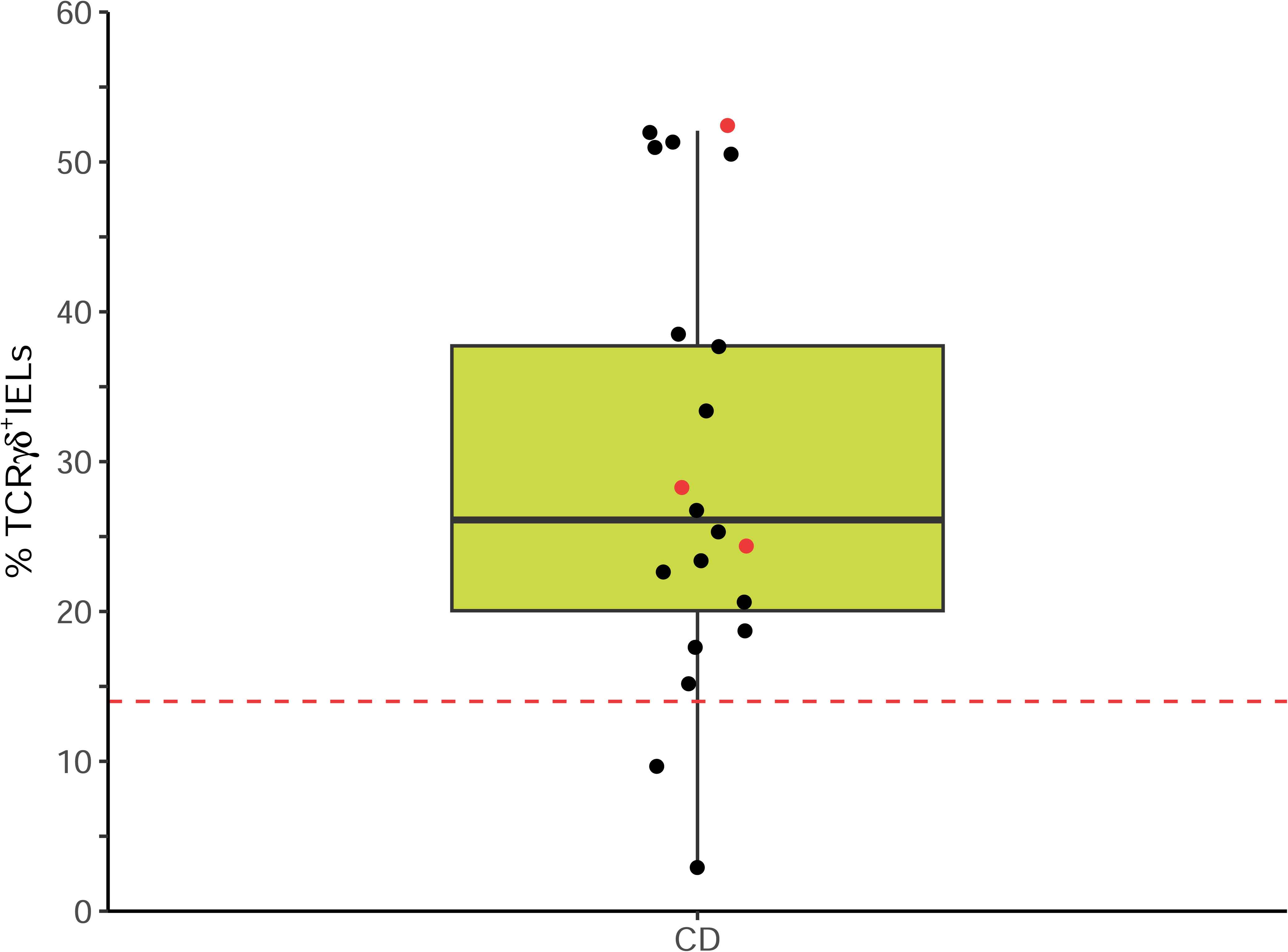
Baseline data of the percentage of TCRγδ^+^ intraepitelial lymphocytes (IELs) in patients with celiac disease (CD). The dotted red line indicates the cut-off for CD diagnosis. Red dots represent subjects with positive fecal GIP.

#### IL-2 levels

All patients in the first subset were analyzed, except for one patient with SGRS. A significant increase when comparing basal and post-4 hour IL-2 levels was observed only within the CD group (p = 7.6*10^-5^) (Figure 4). A positive IL-2 response was observed in 14/17 patients with CD and 2/12 non-CD participants (1 healthy control and 1 patient with SGRS), resulting in 82.4% sensitivity and 83.3% specificity. Only 1 of the 3 patients with CD with positive fecal GIP showed a positive IL-2 response.

**Figure 4.**
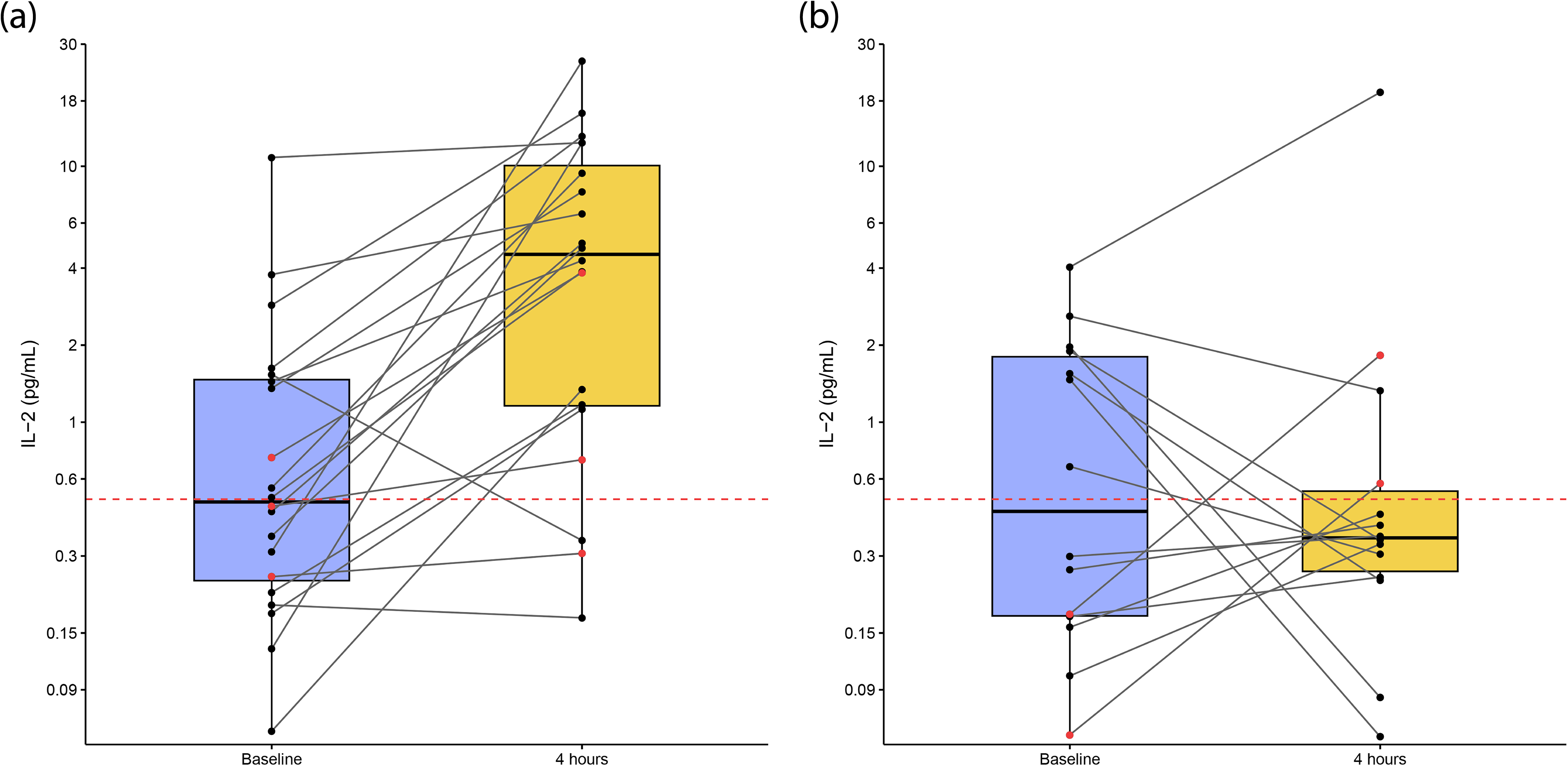
Serum IL-2 data. IL-2 concentration baseline and after 4 hours of gluten challenge is shown in subjects from each study group: (a) patients with celiac disease (CD), and (b) non-CD controls. The dotted red line indicates the cut-off for CD diagnosis. Lines connect results from individual patients. Centerlines in the box show the median; box limits indicate the 25th and 75th percentiles (IQR). Red dots represent subjects with positive fecal GIP.

#### Activated gut-homing CD8^+^ T cells

The calculated AUC was 0.896 (95% CI 0.756-1.036). A cut-off value of 0.010 resulted in 88.2% sensitivity (15/17) and 100% specificity for diagnosing CD (also considering the day 6/day 0 ratio ≥ 2). This threshold is similar to that previously described by our group using sliced white bread for GC [12]. A positive CD8^+^ T-cell response was observed in 2 out of 3 patients with positive fecal GIP.

A significant increase in the CD8^+^ CD103^+^ β7^hi^ CD38^+^ T cell population at day 6 was only observed in patients with CD (p = 2.3*10^-5^) but not in the non-CD control group (Figure 5).

**Figure 5.**
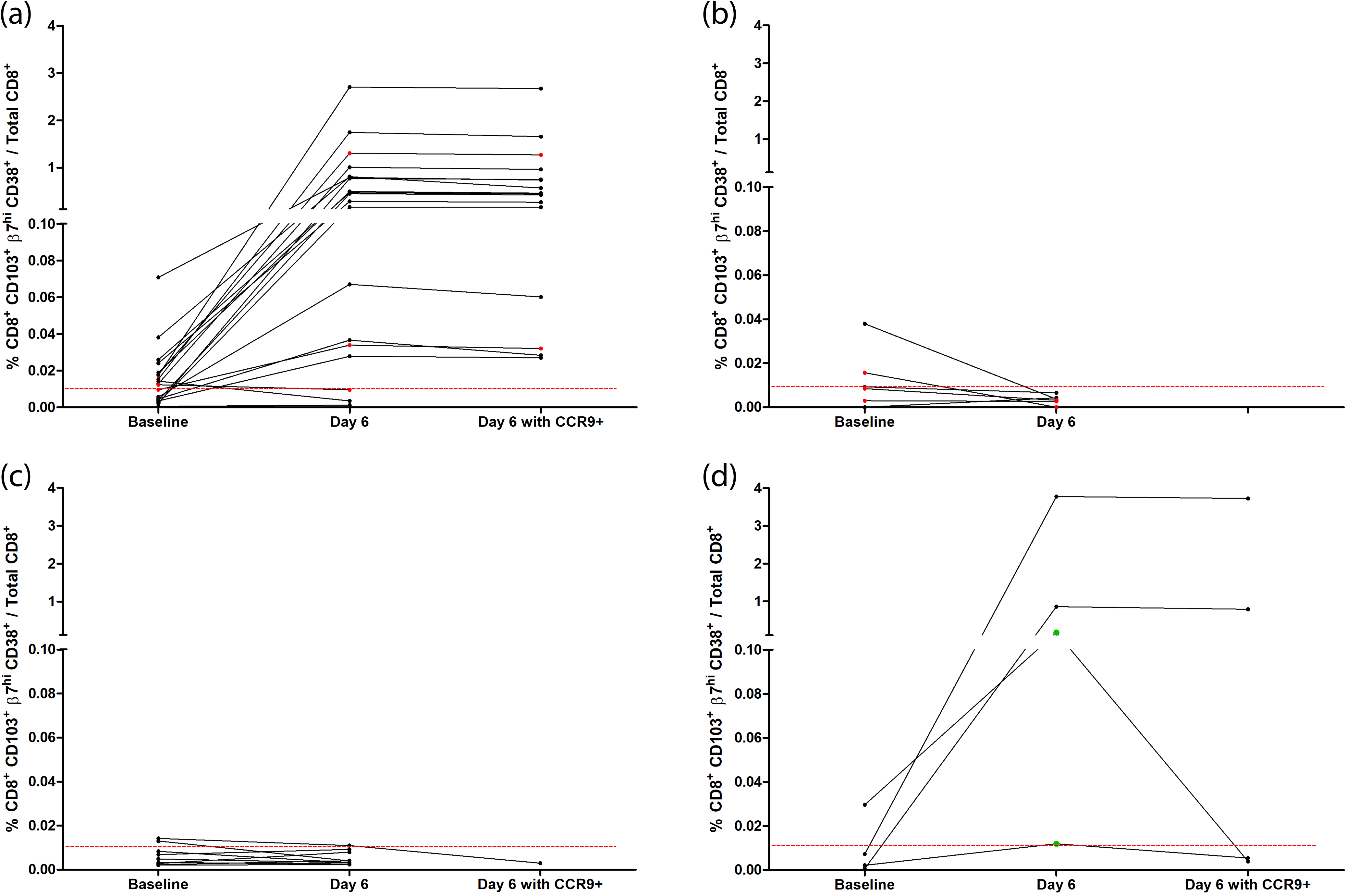
Gut-homing CD8^+^ T-cell responses. Percentage of activated gut-homing CD8^+^ T cells baseline and on day 6 after starting the 3-day gluten challenge is shown in subjects from each study group: (a) patients with celiac disease, (b) patients with suspected gluten-related functional bowel disease symptoms (SGRS), (c) healthy controls, and (d) individuals with uncertain diagnosis. In patients with a response at day 6 exceeding the cut-off (dotted red line), the percentage of CD8^+^ CD103^+^ β7^hi^ CD38^+^ CCR9^+^ cells/total CD8^+^ cells is also shown. Note that including CCR9 eliminates the two false positives (green dots) in the group of uncertain diagnosis. Red dots represent subjects with positive fecal GIP.

In patients with CD, nearly all of the CD8^+^ CD103^+^ β7^hi^ CD38^+^ studied cells also expressed the CCR9 marker at day 6: 96.3% ± 1.3 (range 77.3-98.8), in contrast with the 27.4% observed in the sole healthy control who showed > 0.01% of CD8^+^ CD103^+^
β7^hi^ CD38^+^ cells at day 6. However, this percentage above the established cut-off value of the control cannot be considered a positive CD8^+^ T cell response because the value decreased compared to that observed at baseline.

Ten of the 17 patients with CD showed discrepancies with at least one of the four evaluated diagnostic methods. These differences are presented in Table 2. An increased percentage of TCRγδ^+^ IELS of ≥ 14 was observed in 8 of these 10 patients, which corresponds to the proportion of patients with CD with a positive response to GC for the gut-homing CD8^+^ T cells. However, only one patient tested negative for both assays. IL-2 tested positive in 7 of these 10 patients, all of them positive for the CD8^+^ T-cell assay. According to these results, the percentage of gut-homing CD8^+^ T cells at day 6 showed a borderline significant correlation with the post-4 hour IL-2 levels (ρ = 0.525, p = 0.054) but not with the percentage of TCRγδ^+^ IELs (ρ = 0.226, p = 0.436). A total agreement for increased percentage of TCRγδ^+^, activated gut-homing CD8^+^ T cells, and IL-2 levels was observed in 6 of these 10 patients with CD. *UBE2L3* test provided the lowest sensitivity, as previously mentioned, showing a positive result for the only one CD patient who yielded negative results for the rest of the tests. An increased percentage of TCRγδ^+^ IELs was the only marker present in the three patients with CD with positive fecal GIP results.

**Table 2.** Patients with celiac disease showing a negative response in any of the diagnostic test under study.

| **PATIENT** | **CD8<sup>+</sup> T cells** | **IL-2** | **% TCR$\gamma\delta$<sup>+</sup>** | ***UBE2L3*** |
| --- | --- | --- | --- | --- |
| **Id1** | - | - | - | + |
| **Id2\*** | - | - | + | - |
| **Id3** | - | - | + | + |
| **Id4** | + | - | + | + |
| **Id5** | + | + | - | + |
| **Id6\*** | + | - | + | - |
| **Id7** | + | + | + | - |
| **Id8** | + | + | + | - |
| **Id9** | + | + | + | - |
| **Id10** | + | + | + | - |
| **Id11** | + | + | + | - |
| **Id12** | + | + | + | - |

\* Patients with positive fecal GIP. (-) negative; (+) positive.

#### Symptoms

Clinical symptoms significantly worsened after GC in the CD group (baseline vs. days 1, 2, and 3), with the strongest difference observed between baseline and at post-4 hour (Supplementary Figure 1). Specifically, patients with CD reported exacerbation of abdominal distension, abdominal pain, nausea, tiredness and irritability at 4 hours, but only abdominal distension, abdominal pain, and tiredness persisted on days 2 and 3. Abdominal distension was the only clinical symptom that worsened on day 4.

There was no correlation between clinical symptoms and post-4 hour IL-2 levels, either when all symptoms were considered or when only nausea and vomiting were considered (Supplementary Figure 2).

### Diagnostic utility for CD of the CD8^+^ T-cell assay in patients with uncertain diagnosis: a comparison with the classical extended GC

A total of 45 patients with uncertain diagnosis completed the 3-day and the extended GC.

A CD8^+^ T-cell positive response was observed in 8 patients (Table 3). Their duration of the extended GC ranged from 4 to 20 weeks. In two patients with the shortest GC period (4 and 6 weeks), no serological data could be obtained, but normal mucosa was observed. In another patient who maintained the GC for 6 weeks, negative results for both serology and histology were observed. Notably, the remaining 5 patients showed a GC duration of at least 7 weeks and displayed positive ATG2, although none presented atrophy; however, mucosal lesions intensified with prolonged GC, except for one patient who maintained a GC period of 20 weeks without histological alterations.

**Table 3.**
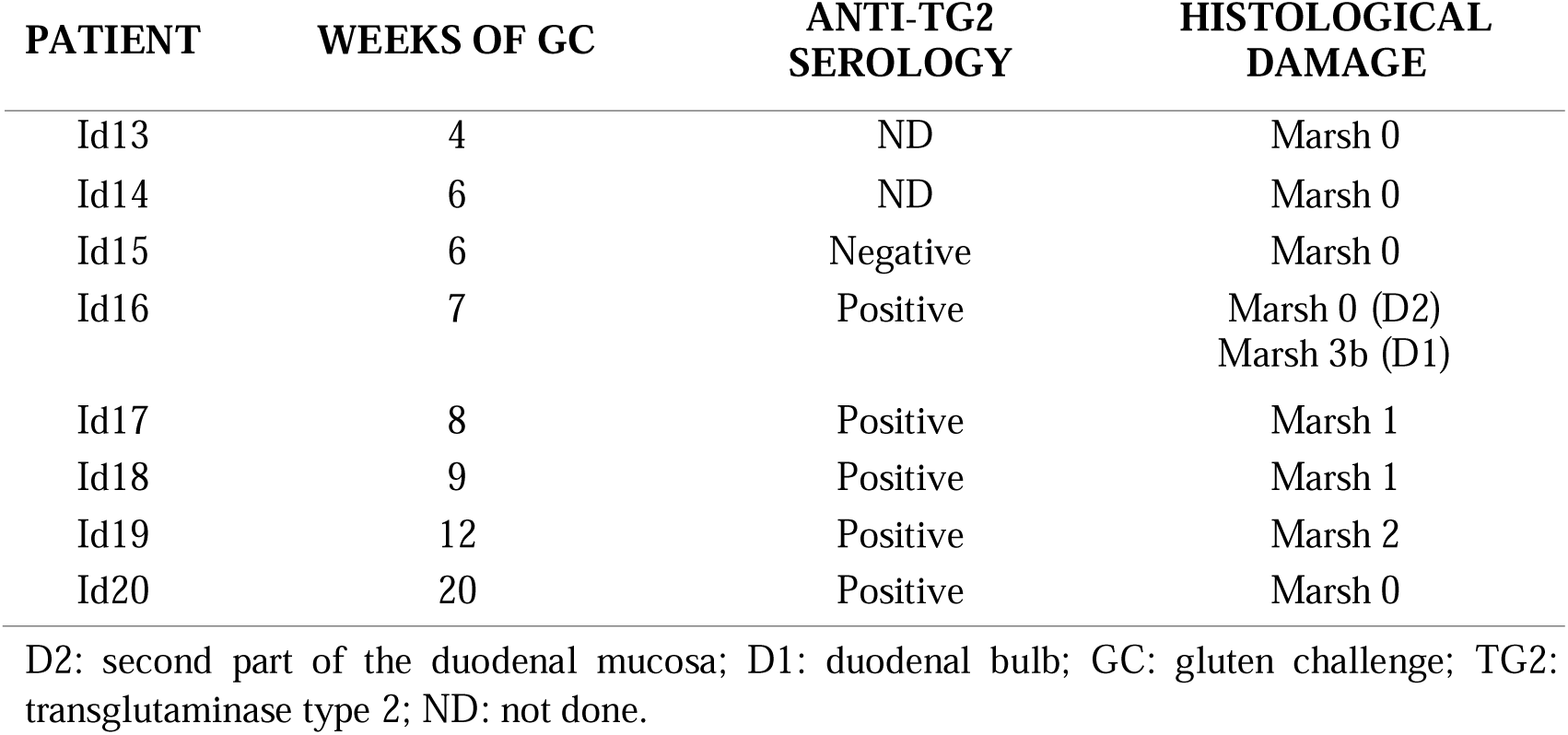
Serological and histological results after extended gluten challenge in patients with a positive response in the CD8^+^ T-cell test.

The CCR9 marker was studied in four patients who showed a positive CD8^+^ T cell response according to the established criteria. In two cases (Id 13 and Id14 of Table 3), a high percentage of the CD8^+^ CD103^+^ β7^hi^ CD38^+^ T cells also presented CCR9^+^, similar to that observed in patients with CD of the first subset. Notably, CD was ruled out in the other two patients after the extended challenge (negative ATG2 and normal mucosa). Less than 40% of the CD8^+^ CD103^+^ β7^hi^ CD38^+^ cells in these two patients were found to be CCR9^+^, which contrasts with the nearly 100% observed in patients with CD (Figure 5).

Furthermore, the 37 patients with a negative CD8^+^ T-cell assay showed negative serology and/or histology after the extended GC, which ranged from 4 weeks to 23 months (median 8 weeks). A total of 33 patients showed normal mucosa (Marsh 0), accompanied by negative ATG2 expression in 28 patients (undetermined for the remainder). One patient presented with Marsh 1 despite negative ATG2. In the remaining three cases, a biopsy was not performed because the patients presented negative serology and non-compatible HLA-genetics.

Clinical symptoms varied greatly among patients, without identifying those with probable CD according to the CD8^+^ T-cell response or ATG2 serology (Supplementary Figure 3).

The data that support the findings of this study are openly available in https://repositoriosaludmadrid.es/ at https://hdl.handle.net/20.500.12530/87937.

## DISCUSSION

Diagnosing CD does not have a universally accepted ’gold standard’. Clinical features, serological markers, or histological findings alone cannot offer a conclusive diagnosis, which requires a comprehensive assessment that combines these factors. Once on a GFD, the complexity increases because of the necessity of implementing a diagnostic algorithm after a GC. The time of response after gluten reintroduction is not uniform among all individuals, making it challenging to establish a balance between short-term gluten exposure for symptom relief and achieving serological and histological responses. In contrast, alternative methodological approaches that provide more consistent responses across individuals have been described in recent years. Interestingly, they do not require GC [13, 15, 16], or reduce it to only one [19] or three [7, 8, 9] doses.

In the present study, we have compared the diagnostic accuracy of four previously reported methods for the diagnosis of CD in individuals on a GFD. A similar sensitivity was found for the percentage of TCRγδ^+^ IELs and the CD8^+^ T-cell assay (88.2%), with slightly lower sensitivity noted for the IL-2 test (82.4%). The *UBE2L3*-based test showed low sensitivity (52.9%) despite maximum specificity.

The analysis of the percentage of TCRγδ^+^ IELs does not require GC, potentially offering a significant advantage by making it applicable to individuals unwilling to undergo gluten reintroduction or in scenarios where GC is contraindicated (e.g., anaphylactic reactions to gluten or severe neurological manifestations). Additionally, this method is not affected by dietary transgressions, as it can be used in the diagnostic work-up of patients on a gluten-containing diet [14]. This may increase its sensitivity compared to other proposed methods, as indicated by the findings in patients with positive fecal GIP. One possible limitation of this approach, which could not be addressed in this study, is its specificity. Although the so termed “celiac lymphogram”, which is characterized by an increase in the TCRγδ^+^ IEL subset and a decrease in the CD3^−^ IEL subset, provides the highest accuracy for CD diagnosis for active disease, the CD3^−^ IEL subset tends to recover with mucosal healing and can reach normal values after gluten withdrawal [14]. However, specificity is still maximal in patients who maintain low CD3^-^ on a GFD. Remarkably, increased percentages of TCRγδ^+^ IELs have been described as useful in the work-up and management of seronegative villous atrophy [25, 26] and in the differential diagnosis of CD and non-celiac gluten sensitivity [15].

Methods that require GC eliminate the need for the invasive procedure to obtain duodenal samples. The test based on activated gut-homing CD8^+^ T cells yielded 88% sensitivity and 100% specificity. We have previously described a 95% specificity for this methodology [12]. However, the present study observed that the addition of the CCR9 marker seems to rule out a positive CD8^+^ T-cell response in some complex patients showing low positive CD8^+^ T-cell results. Similarly, low responses were observed in the individuals who had previously demonstrated a false-positive result. Possibly, they would have demonstrated an accurate result using this new configuration. Further studies are necessary to confirm whether a specificity higher than 95% is expected in this assay when CCR9 is included. Interestingly, CCR9 has been described to characterize the phenotypic profile of gluten[specific CD4^+^ T cells and it seems to be present also in gluten-induced CD8^+^ T cells [27]. The IL-2 test showed 82% sensitivity and 83% specificity. The difference in sensitivity between the two assays was based on a single patient who was detectable only by the CD8^+^ T cell assay. In contrast, 100% specificity has been previously reported for IL-2 [19]. A difference from the previous study is that healthy unaffected individuals consuming an unrestricted diet were used as controls. Given the limited number of controls included in the studies conducted thus far, it is plausible that the difference between our results and those previously reported stems from the specific characteristics of the individuals studied.

Beyond these differences in diagnostic accuracy, the CD8^+^ T-cell-based assay has additional advantages. The only technical requirement is a flow cytometer, which is used in clinical settings and readily available in most hospitals. Each sample could be handled in fresh and individually, yielding the results for each patient within approximately 2 hours after collecting the second blood sample (day 6). Furthermore, samples can be also processed within 24 hours of collection, enabling their transfer to alternative centers for analysis. In contrast, IL-2 is present at very low concentrations in the blood, requiring the use of high-sensitivity platforms for analysis. Such platforms are rare and are typically confined to research environments. Additionally, to ensure cost-effectiveness, samples are not processed individually, but are held until a certain number are collected, causing delays in obtaining results. Finally, despite the lack of an observed correlation between IL-2 levels and symptoms in our study, this correlation has been documented, which may potentially reduce the sensitivity of the analysis in certain patients. An advantage that can be ascribed to the IL-2 assay is that it needs a single dose of gluten in contrast to the three doses required by the CD8^+^ T-cell based test. Nevertheless, upon closer examination of symptomatology, it becomes apparent that the greatest clinical worsening is observed after the first dose of gluten, although it should be noted that this may vary from patient to patient.

The *UBE2L3* assay requires further attention. It also circumvents the need for GC, and has potential as a complementary tool to the aforementioned approaches, given its ability to yield a positive result in cases where false negatives are encountered with the other studied assays. Its main limitation is low sensitivity; however, it is a minimally invasive tool with high specificity, eliminating the need for gluten reintroduction. *UBE2L3* has emerged as a promising diagnostic marker following a Mendelian randomization analysis using publicly available GWAS, expression QTL (eQTL), and methylation QTL data [16], and subsequent validation using independent expression data of PBMCs from patients with CD. Of particular interest is the association of *UBE2L3* genetic variants with susceptibility to various autoimmune disorders, including CD, as well as the UBE2L3 involvement in NF-κB activation, a pathway implicated in CD [28]. This is the first prospective study considering *UBE2L3*; however, the aforementioned findings warrant large-scale investigations to establish definitive conclusions regarding its potential use as a CD biomarker.

The present study also aimed to demonstrate the utility and feasibility of the CD8^+^ T-cell assay as a diagnostic tool in the clinical settings. A total of 45 patients underwent a 3-day GC followed by a more standard GC (range: 4-92 weeks, median 8 weeks). Variability in the duration of gluten exposure reflects real-world clinical scenarios. Although a minimum of 6-8 weeks of gluten consumption was recommended, some patients declined or discontinued due to clinical discomfort. Throughout the study period, we observed that a few patients refused the 3-day GC, but the proportion greatly increased when longer challenges were proposed. Therefore, there is an evident need for a short-term GC in clinical practice. In these 45 patients, CD diagnosis was based on the final serology and biopsy results according to the current guidelines. However, atrophy was not detected in any of these patients. Five patients with a positive CD8^+^ T-cell response had positive serology with minimal mucosal changes. The combined serological and cell responses provide compelling evidence supporting the diagnosis of CD and indicate that 6-8 weeks may not be sufficient to provide a definitive diagnosis in all patients, especially if gluten intake was low. Unfortunately, the other two patients who also showed a strong CD8^+^ T-cell response (Id13 and Id14) failed to attend the serological evaluation after GC. However, both presented HLA-DQ2.5 and a celiac lymphogram despite having normal mucosa. Notably, they followed a diet low in gluten for 4 and 6 weeks, respectively, because of clinical discomfort and one of them had a positive ATG2 result in a previous analysis while on a gluten-containing diet.

This pilot study conducted in clinical practice demonstrated the feasibility of integrating the assessment of activated gut-homing CD8^+^ T cells into clinical protocols. Moreover, this study sheds light on the diverse scenarios in which this diagnostic approach can prove beneficial. While individuals following a self-prescribed GFD or those with incomplete testing at onset (on a gluten-containing diet) are obvious candidates, our study revealed its utility in complex cases, such as those showing low ATG2 levels. ATG2 is the most sensitive serological test used for diagnosing CD; however, its specificity may be diminished, particularly at lower titers [29, 30, 31]. The isolated presence of ATG2 serology cannot be considered CD-specific. The same is true for histology: seronegative villous atrophy is a clinical challenge. Importantly, the present work also demonstrates that the CD8^+^ T-cell assay can show higher accuracy for diagnosing CD than current standard practices in cases of uncertain diagnosis, as it offers more consistent responses across patients after gluten reintroduction.

The kinetics of serological and histological responses to GC likely depend on various factors, resulting in significant heterogeneity among individuals. Prolonged GC is necessary to induce atrophy in patients with CD; however, the emergence of clinical symptoms often deters many from continuing gluten intake. Moreover, studies assessing the duration and quantity of GC mostly included patients with high ATG2 levels and initial villous atrophy. However, individuals with initially milder enteropathy, often accompanied by lower ATG2 levels, may require extended periods of gluten exposure for an accurate diagnosis. The assays presented herein capture early changes induced by a short-term gluten reintroduction, yielding a consistent and uniform response across subjects, including differences observed despite initiating a GFD (increased TCRγδ^+^ IELs). The feasibility of the intraepithelial lymphogram in routine clinical practice has been already proved [13, 14]. Furthermore, we demonstrated the clinical applicability of the CD8^+^ T-cell assay. These tools offer novel approaches to facilitate diagnosis in individuals on a GFD, which can be employed individually or in combination, depending on the clinical context or individual circumstances.

The absence of a standardized GC protocol also affects children, making them potential beneficiaries of the findings outlined here. Recently, a clinician’s guide for GC in children was published, recommending a minimum intake of 3–6 g of gluten per day for at least 12 weeks to enhance the accuracy of CD diagnosis [32]. It is especially relevant that GC is discouraged in children under five years of age or during pubertal development [5]. However, a shorter 3-day GC or an upper endoscopy with biopsy could be conducted during this period, presenting a significant advantage.

Currently, accurate diagnosis of CD remains a persistent challenge. Achieving a precise diagnosis is crucial for effective treatment and prevention of life-threatening complications, and also for mitigating the costs associated with inappropriate testing, incorrect care, and potential malpractice claims. Equally significant is the social, psychological, and economic impact of incorrectly prescribing a GFD to individuals who do not have CD or other gluten-related disorders. This study represents a significant advancement in improving the diagnosis of CD by introducing tools such as the intraepithelial lymphogram and CD8^+^ T-cell assay, which are easily implementable in clinical practice.

## Abbreviations used in this paper

CD: celiac disease
CI: confidence interval
GC: gluten challenge
GFD: gluten-free diet
GIP: gluten immunogenic peptides
IELs: intraepithelial lymphocytes
TG2: transglutaminase type 2.

## Data Availability

Main data that support the findings of this study are openly available in https://repositoriosaludmadrid.es/ at https://hdl.handle.net/20.500.12530/87937.

## ACKNOWLEDGEMENTS

We thank the collaboration of the institutions and participant centers for volunteer recruitment and the generous volunteers who enrolled in the study.

---

**Supplementary Figure 1.** Visual Analog Scale (VAS) scores of celiac disease patients baseline and during the 6 days of study. Individual scores were obtained by summing the responses for flatulence, abdominal distension, abdominal pain, diarrhea, nausea, vomiting, tiredness, irritability and brain fog, ranging from 9 to 45 (representing the absence of symptoms or the presence of severe symptoms in the 9 recorded items).

**Supplementary Figure 2.** Interleukin-2 serum concentration at 4 hours after the first dose of gluten in relation to symptoms severity as measured by (a) total Average Visual Analog Scale (VAS) score or (b) nausea and vomiting VAS score.

**Supplementary Figure 3.** Average Visual Analog Scale (VAS) score in the group of patients with uncertain diagnosis. VAS score was calculated considering the 6 days of study after the GC for each recorded symptom. Red dots indicate patients showing a positive CD8^+^ T-cell response to the 3-day gluten challenge. Note that these data correspond to 24 patients.

