## Supplementary figures and images for "Diagnosis of celiac disease on a gluten-free diet: a multicenter prospective quasi-experimental clinical study"

### Supplementary Figure 1

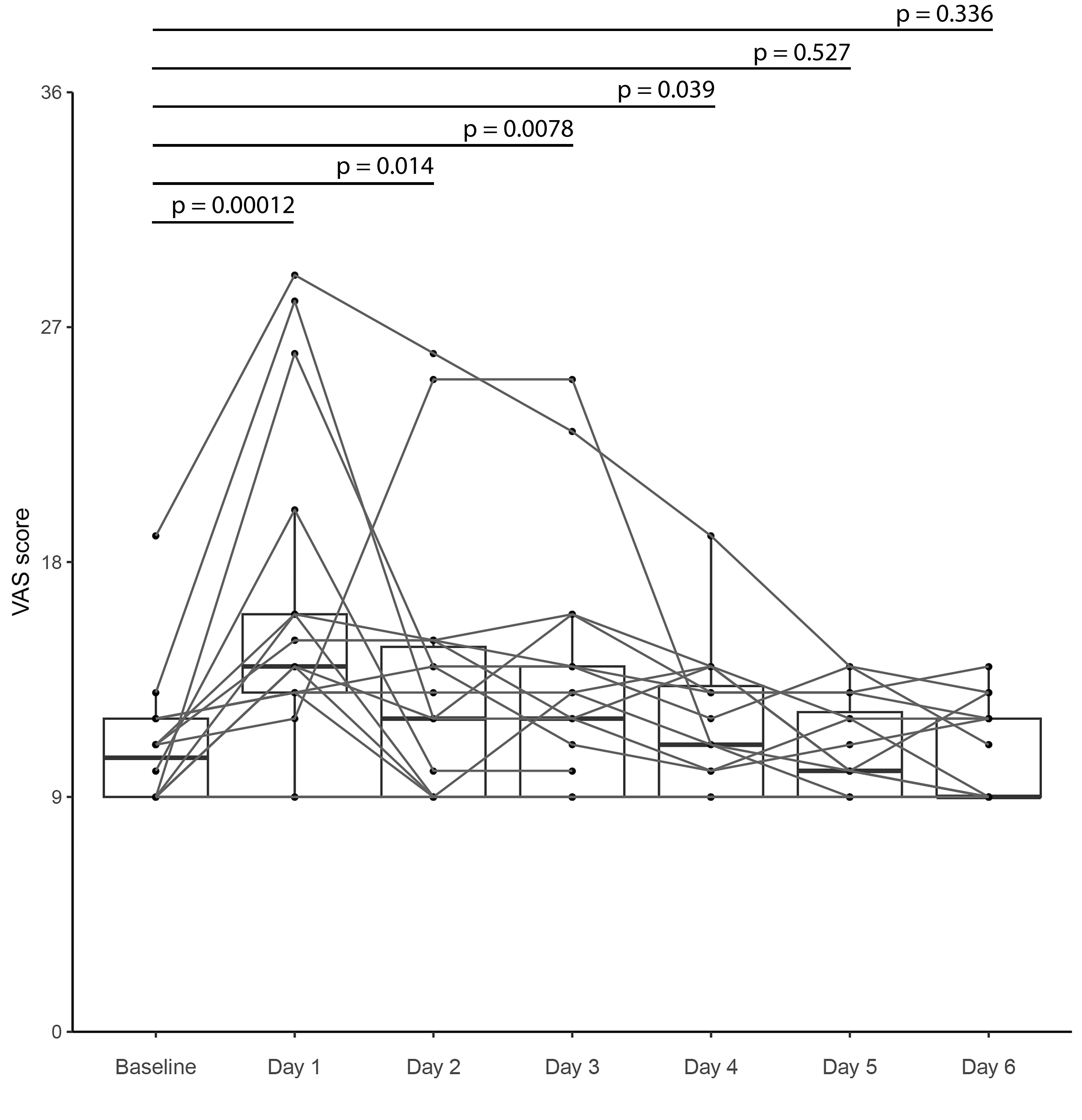

### Supplementary Figure 2

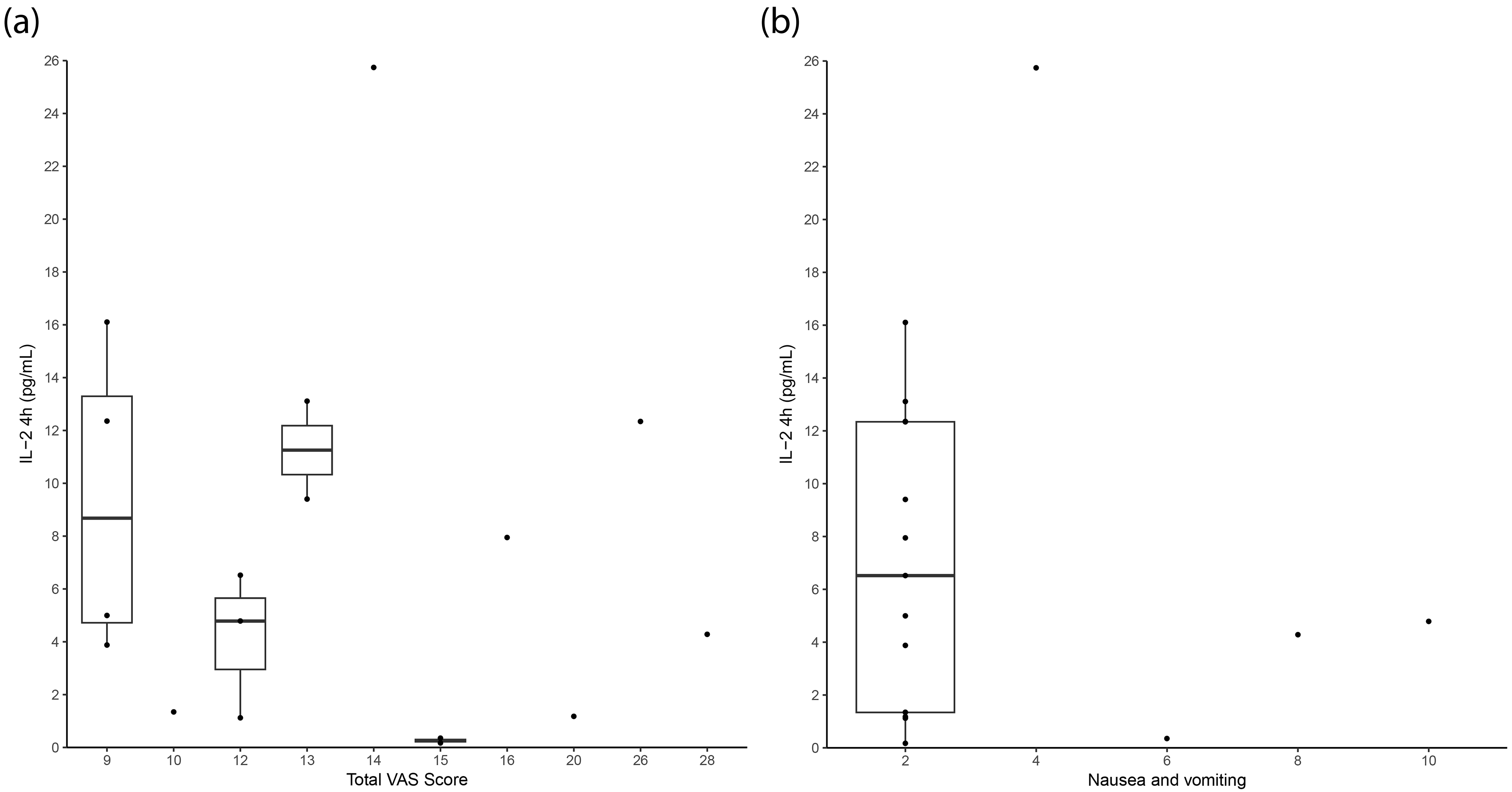

### Supplementary Figure 3

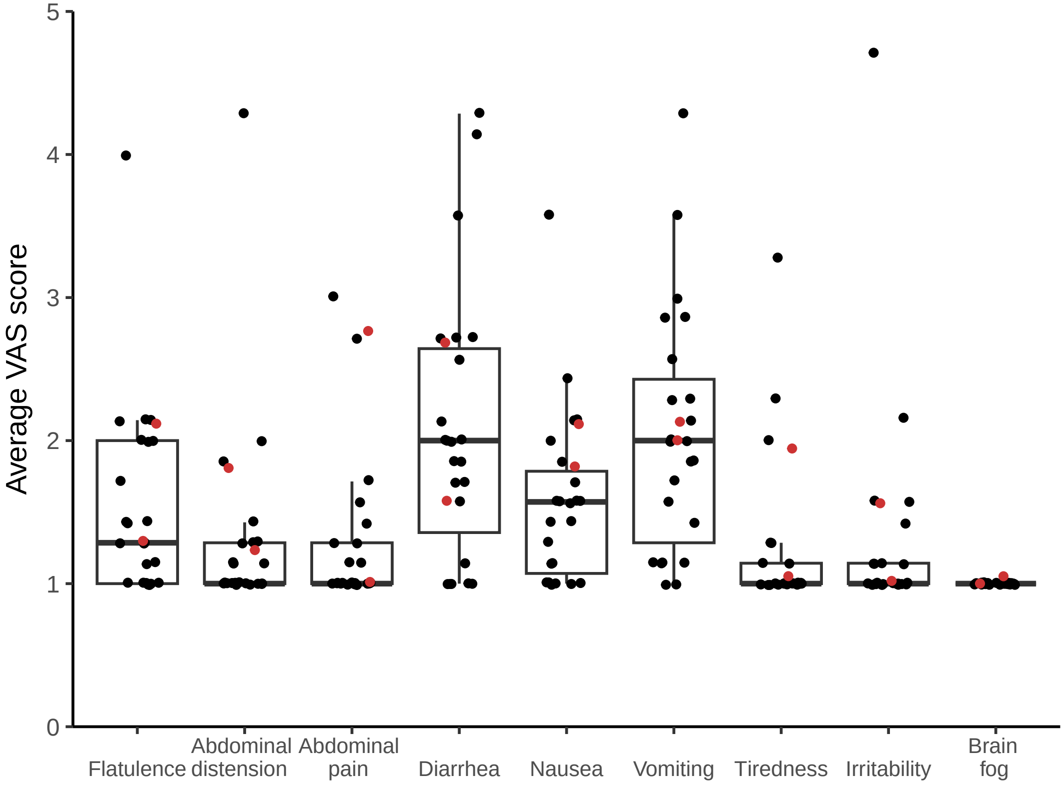
